# Pregnancy Reduces COVID-19 Vaccine Immunity Against Novel Variants

**DOI:** 10.1101/2025.01.29.25321357

**Authors:** Maclaine A. Parish, Jaiprasath Sachithanandham, Lizeth Gutierrez, Han-Sol Park, Anna Yin, Katerina Roznik, Patrick Creisher, John S. Lee, Laura A. St Clair, Annie Werner, Catherine Pilgrim-Grayson, Lea Berhane, Hana Golding, Patrick Shea, Katherine Fenstermacher, Richard Rothman, Irina Burd, Jeanne Sheffield, Andrea L. Cox, Andrew Pekosz, Sabra L. Klein

## Abstract

Pregnant women are at heightened risk for severe outcomes from infectious diseases like COVID-19, yet were not included in initial vaccine trials, which may contribute to low booster uptake (15% or lower). We explored the serological and cellular responses to COVID-19 mRNA booster vaccines (i.e., ancestral and BA.5) in pregnant and age-matched, non-pregnant females to identify how pregnancy affects immunity against vaccine and novel variants. Antibodies from pregnant women were less cross-reactive to non-vaccine antigens, including XBB.1.5 and JN.1. Non-pregnant females showed greater IgG1:IgG3 ratios and neutralization against all variants. In contrast, pregnant women had lower IgG1:IgG3 ratios and neutralization but increased antibody-dependent NK cell cytokine production and neutrophil phagocytosis, especially against novel variants. Pregnancy increased memory CD4+ T cells IFNγ production, monofunctional dominance, and fatty acid oxidation. Pregnancy may reduce the breadth, composition, and magnitude of humoral and cellular immunity, particularly in response to novel variants.

## Introduction

Pregnant women faced significant morbidity and mortality during the COVID-19 pandemic, with an elevated risk for severe outcomes [1–3]. From the onset of the pandemic in early 2020 to December 2022, there were 24,203 cases of COVID-19 among pregnant women in the United States with 2,043 fatalities [4]. This 8.4% mortality rate is significantly higher than that of the general population (0.061%), reflecting the elevated risk for disease outcomes in pregnancy. Pregnant women also were more likely to require hospitalization and invasive ventilation from acute SARS-CoV-2 infection than non-pregnant women [3, 4]. Adverse pregnancy and neonatal outcomes were an additional concern, as SARS-CoV-2 infection has been associated with preeclampsia, stillbirth, NICU admission, and preterm birth in women [1, 5–9] and mouse models [10].

American College of Obstetricians and Gynecologists (ACOG) recommend vaccination during pregnancy to provide optimal protection against SARS-CoV-2 for both the mother and fetus [11]. Data from November 2021 show vaccination rates among pregnant women (45.1%) were significantly lower than that of non-pregnant individuals (64.9%) [12]. Vaccination rates among pregnant women have since fallen precipitously, and remain lower than the general population, with less than 15% of pregnant women vaccinated in the 2023-2024 vaccination season [13]. This decline in vaccine uptake is particularly concerning given that other vaccines offered during pregnancy, such as influenza and Tdap, show higher coverage rates (47.4% and 59.6%, respectively) [14, 15]. Despite a favorable benefit of administering COVID-19 vaccines during pregnancy [16–18], low vaccine uptake may be a result of greater vaccine hesitancy in this population [19–24].

While vaccination is the best preventative strategy to reduce the severity of COVID-19 outcomes [16, 25], existing evidence suggests pregnant women mount dampened immune responses to vaccination, highlighting the need for deeper immunological assessments [26]. While evidence investigating the immunogenicity of COVID-19 vaccines in pregnant women showed effective neutralizing antibody responses to primary mRNA vaccine two-shot series and efficient maternal antibody transfer via cord blood [27–29], data on cross-protection elicited by vaccination are limited. Previous studies of COVID-19 vaccines in pregnant women have focused on placental antibody transfer, demonstrating the benefits of maternal vaccination in protecting newborns and infants from severe infection [30–32]. While several studies have aimed to compare the antibody landscape induced by vaccination during pregnancy with those in non-pregnant women [33], to date, the effects of pregnancy on cross-protection against circulating variants in addition to vaccine antigens have remained unexplored.

Using systems serology, we can broadly and deeply characterize the antibody landscape in response to COVID-19 vaccination. By employing this systematic and comprehensive approach, we can gain insight to shape future vaccine development and public-health campaigns by understanding the protective immune response generated by vaccination [34]. Combining functional assays and systems serology methods, we show that the breadth and diversity of antibody generated by vaccination and polyfunctionality of spike-specific memory CD4+ T cells are reduced during pregnancy.

## Results

### Pregnancy alters IgG subtype dominance in response to novel variants

Because SARS-CoV-2 nucleocapsid (N) antibodies are not generated by mRNA vaccines targeted to S protein, this can be used as a proxy for SARS-CoV-2 infection [46], with recognized limitations [37]. There was no increase in either anti-N IgG **(Figure 1A)** or in any subclass of IgG1-4 **(Supplemental Figure 1A)** in any participants, suggesting that no one was infected with SARS-CoV-2 during their study enrollment window. However, all participants had detectable anti-N IgG levels highlighting prior exposure to SARS-CoV-2 in our cohort. Luminex-bead assays were used to comprehensively characterize the antibody response generated by vaccination against SARS-CoV-2 vaccine antigens (ancestral and BA.5 S) and circulating variants (XBB.1.5 and JN.1 S). COVID-19 boosters induced IgG in both pregnant and non-pregnant females to circulating variants **(Figure 1B)**. Similar to anti-N IgG, prior to vaccination, total IgG to ancestral S and BA.5 S was high in both cohorts, as SARS-CoV-2 exposure through infection and prior vaccination is pervasive in the general population [47]. This was reflected by limited increase in anti-S IgG responses to vaccine variants (ancestral and BA.5) as pre-existing immunity was high at baseline. In contrast, anti-S IgG increased in response to novel variants (XBB.1.5 and JN.1) likely as a result of lower pre-existing immunity. There were, however, distinct differences in the IgG subtype dominance across groups. When comparing relative isotype abundance, there was IgG1 subtype dominance among non-pregnant, but not pregnant, females, particularly in response to JN.1 (p = 0.04) **(Figure 1C-F).** While IgG3 contributed to a small proportion of total IgG generated in response to vaccination, pregnant women were more likely to induce anti-S IgG3 than non-pregnant females, particularly against BA.5, while non-pregnant women were more likely to induce IgG4 **(Supplemental Figure 1 B-E**). Non-pregnant women induced anti-S IgG4 responses against all variants. While this is atypical for traditional vaccine responses, evidence in the literature suggest this may be a signature of immune imprinting from subsequent vaccination with mRNA boosters [48]. While not investigated here, it has been suggested that this may be evidence of immune tolerance and may be related to incidence of breakthrough infection [48, 49].

**Figure 1.**
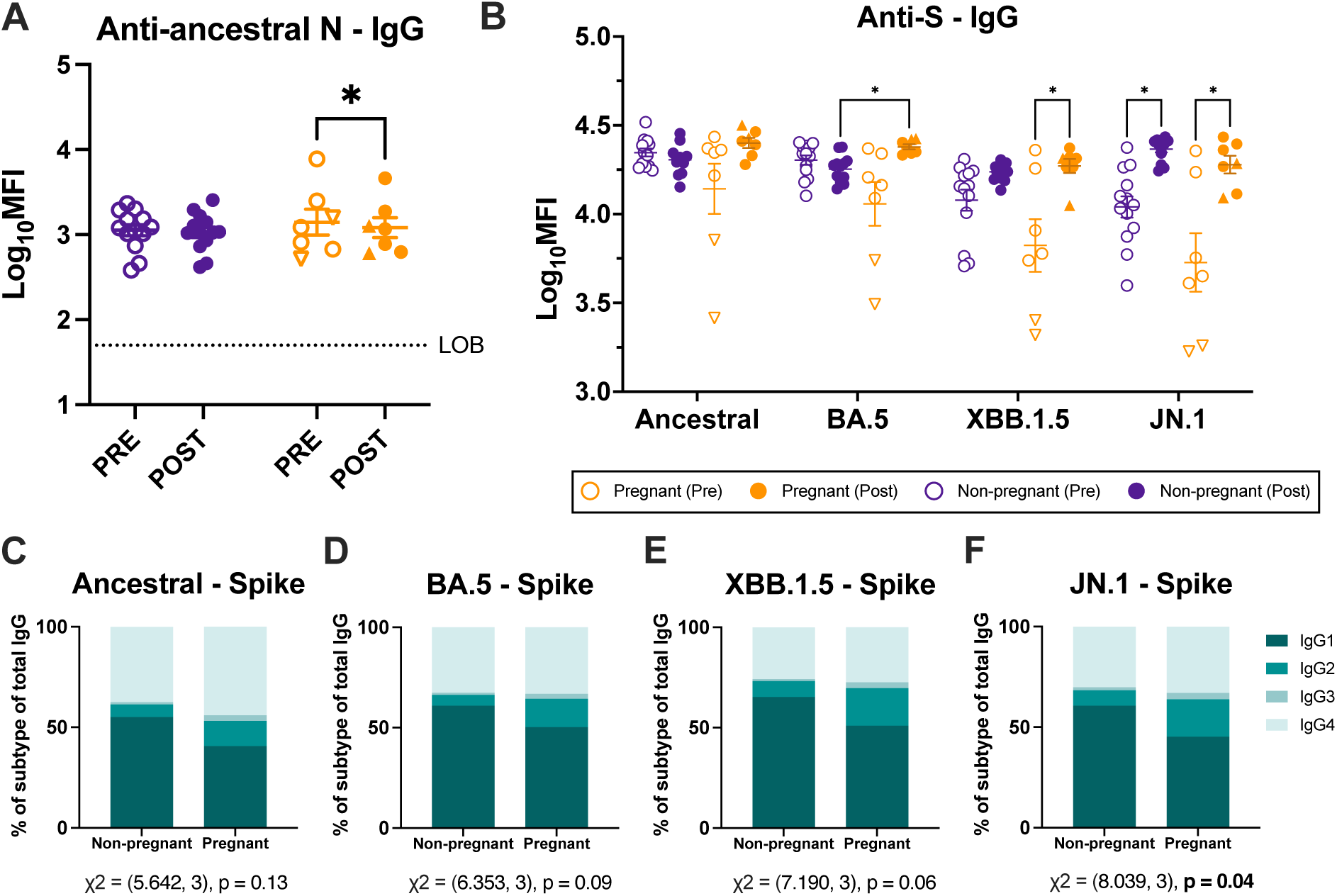
Vaccination differently induces IgG subtype responses during pregnancy. IgG antibody responses in serum pre and 3-5 weeks post-vaccination were measured by systems serology and reported as log_10_ mean fluorescence intensity. Pregnant women (n=7) are shown in orange and non-pregnant females (n=13) in purple. Triangles highlight women who received monovalent boosters and circles represent bivalent booster recipients. Horizontal bars indicate mean MFI across samples with bars indicating standard error of the mean. **(A)** Total IgG to ancestral nucleocapsid (N) protein**. (B)** Total IgG separated by SARS-CoV-2 vaccine (ancestral and BA.5) and variant (XBB1.5 and JN.1) spike (S) antigen and pregnancy status. **(C-F)** Relative abundance of IgG subtypes in pregnant and non-pregnant women as stacked bar plots representing each isotype as a percentage of Total IgG by **(C)** ancestral S, **(D)** BA.5 S, **(E)** XBB.1.5 S, and **(F)** JN.1 S. Data were analyzed by (A) repeated measures two-way ANOVA (B) linear mixed effects regression analysis with Bonferroni post-hoc. (C-F) Isotype proportions were analyzed by Chi-squared analysis with values reported under each corresponding graph. Asterisks indicate p< 0.05. Stippled line indicates limit of background (LOB) defined as the log_10_MFI of a defined negative control.

### Pregnancy reduces the cross-neutralizing capacity of antibodies recognizing SARS-CoV-2 variants

The ratio of IgG1:IgG3 has been shown to be predictive of SARS-CoV-2 neutralization capacity [50]. Compared with non-pregnant females, pregnant women had reduced IgG1:IgG3 ratios, particularly to novel variants, (JN.1 p = 0.02, XBB.1.5 p = 0.05) suggesting that the dominant IgG1 response in non-pregnant females may be indicative of broader cross-neutralization **(Figure 2A-D).** This was reflected by reduced neutralizing antibody (nAb) titers in pregnant compared with non-pregnant women to variants (JN.1 p = 0.07, XBB.1.5 p = 0.03). Both cohorts showed a rise (p < 0.001) in nAb titers to ancestral and BA.5 viruses after vaccination and were able to effectively neutralize these viruses **(Figure 2E-F)**. Pregnant women, however, had significantly lower nAb titers than non-pregnant females, with many pregnant women failing to reach the limit of detection **(Figure 2E-H)**. In response to XBB.1.5, while vaccination significantly increased nAb titers in both groups, these responses were significantly higher in non-pregnant than pregnant females, with 43% (3/7) of pregnant women failing to reach detectable nAb titers in comparison to 15% (2/13) of non-pregnant women. Likewise, among non-pregnant females, vaccination boosted nAb titers against JN.1; there was, however, no vaccine-induced increase in nAb titers among pregnant women, with only 28% (2/7) surpassing the limit of detection. **(Figure 2G-H)**. This is further reflected by modest positive correlations between neutralizing antibody titers and IgG1/IgG3 ratios, among non-pregnant women **(Figure 2J-L),** suggesting their increased IgG1/IgG3 ratios may contribute to the increased neutralizing capacity in comparison to pregnant females, even with small sample sizes. These data suggest that there is reduced breadth and cross-reactivity to variants after COVID-19 boosting among pregnant compared with non-pregnant females. A reduction in neutralizing antibody titer, potentially facilitated by a shift in isotype dominance may contribute to reduced protection from vaccination during pregnancy.

**Figure 2.**
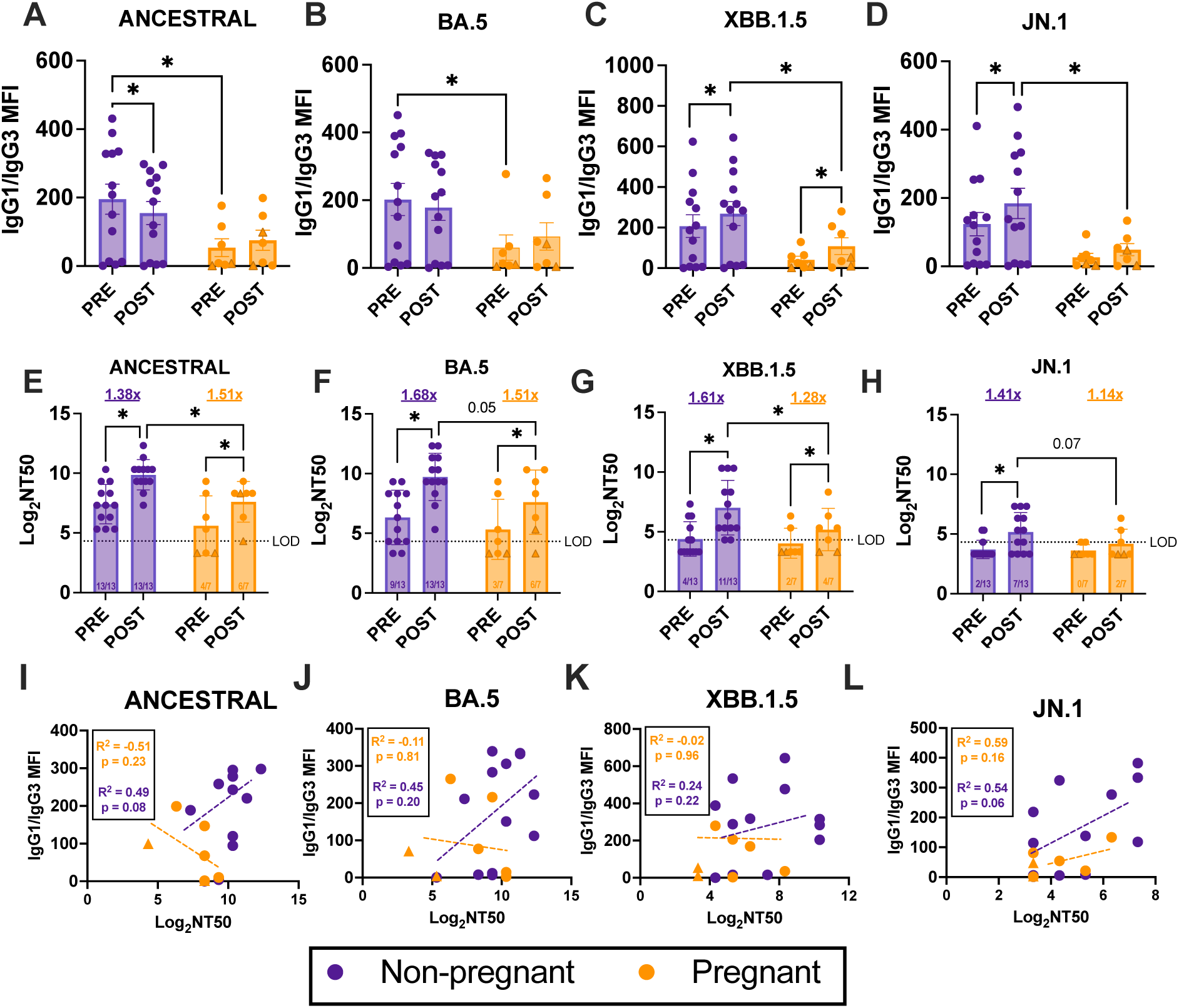
Pregnancy alters cross-neutralization of variants of concern. IgG antibody responses in serum pre and 3-5 weeks post-vaccination were measured by systems serology and reported as mean fluorescence intensity. Pregnant women (n=7) are shown in orange and non-pregnant females (n=13) in purple. Triangles highlight women who received monovalent boosters and circles represent bivalent booster recipients. Bars represent mean MFI across samples with bars indicating standard error of the mean. **(A-E)** Ratio of IgG1:IgG3 antibodies specific to **(A)** ancestral spike (S), **(B)** BA.5 S, **(C)** XBB.1.5 S, and **(D)** JN.1 S. Neutralizing antibody titers reported as Log_2_ NT50 against **(E)** ancestral, **(F)** BA.5, **(G)** XBB.1.5, and **(H)** JN.1 viruses. Ratios shown on the x-axis indicate number of women per group/timepoint that are above the limit of detection (LOD) indicated by the stippled line. **(I-L)** Pearson correlations between IgG1:IgG3 ratios and neutralizing antibody titers reported as Log_2_ NT50 to **(I)** ancestral spike (S), **(J)** BA.5 S, **(K)** XBB.1.5 S, and **(L)** JN.1 S. (A-H) Data were analyzed by repeated measures two-way ANOVA with Bonferroni post-hoc. Asterisks indicate p< 0.05. (I-L) Regression lines for each group are shown in dashed lines colored according to cohort. Separate R^2^ and p-values are shown for each variant separated by pregnancy status.

### Fc receptor binding is not increased in pregnant women against novel variants

While nAb titers are considered a correlate of protection for SARS-CoV-2 infection [51–53], non-neutralizing functions are also important and may contribute to robust protection from severe disease [54]. Data from mouse models suggest FcR-binding and its associated effector functions are necessary for preventing severe COVID-19 disease [55]. To further understand the depth and breadth of antibody generated by mRNA boosters in pregnant women, we investigated FcR binding. Antibody Fc binding to FcγRIIA-R, FcγRIIA-H, FcγRIIb, FcγRIIIA-V, FcγRIIIA-F, FcγRIIIB, FcRn, and FcαR was assessed by a multiplexed Luminex bead assay. FcR binding was similarly induced by vaccination in response to ancestral and BA.5 vaccine antigens among both non-pregnant and pregnant females **(Figure 3A-B).** While non-pregnant females showed similar increases in FcR binding to non-vaccine antigens following vaccination, vaccination did not induce robust changes in FcR binding in pregnant women, with increased binding to only 2/6 FcγRs in response to XBB1.5 and 4/6 FcγRs in response to JN.1 (**Figure 3C-D**).

**Figure 3.**
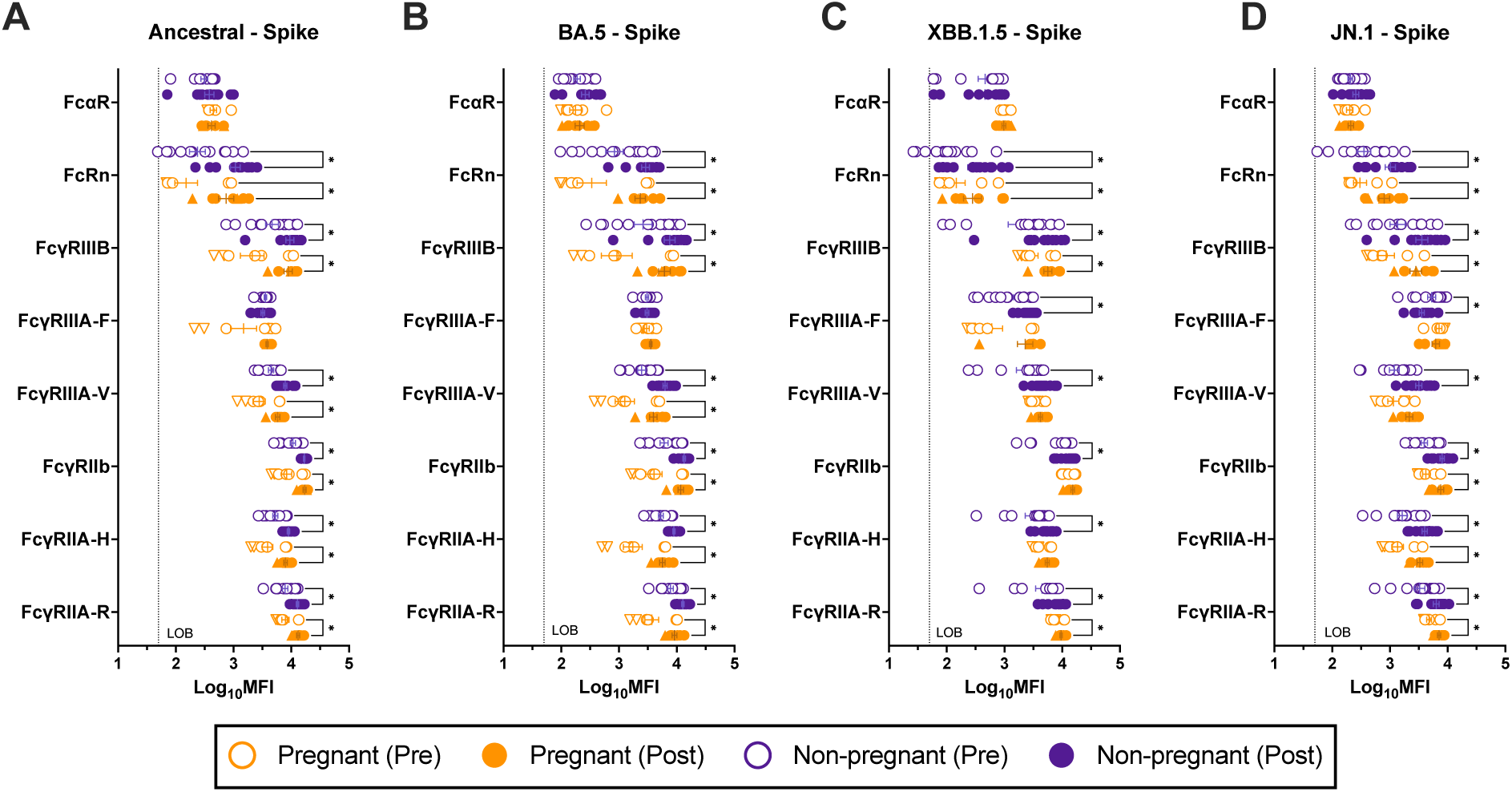
Fc receptor binding is reduced in pregnant women in response to variants of concern. Serum antibody Fc receptor (FcR) binding pre and 3-5 weeks post-vaccination were measured by multiplex Luminex bead assays and reported as log_10_ mean fluorescence intensity. Pregnant women (n=7) are shown in orange and non-pregnant females (n=13) in purple. Triangles highlight women who received monovalent boosters and circles represent bivalent booster recipients. Horizontal bars indicate mean MFI across samples with bars indicating standard error of the mean. Antibody binging to FcγRIIA-R, FcγRIIA-H, FcγRIIb, FcγRIIIA-V, FcγRIIIA-F, FcγRIIIB, FcRn, and FcαR was determined for **(A)** ancestral spike (S), **(B)** BA.5 S, **(C)** XBB.1.5 S, and **(D)** JN.1 S. Data were analyzed by linear mixed effects regression analysis with Bonferroni post-hoc. Asterisks indicate p< 0.05. Stippled line indicates limit of background (LOB) defined as the log_10_MFI of a defined negative control.

### Non-neutralizing antibody functions and nAb are negatively correlated among pregnant women

To understand how FcR binding contributes to immunity generated by COVID-19 vaccination, several non-neutralizing antibody functions were assessed. Non-neutralizing functions aid in the clearance of infected cells through recruitment of effector cells [56]. Antibody-dependent complement deposition (ADCD) was analyzed with a multiplexed Luminex bead assay to measure antibody binding to complement protein C3 as a broad indicator of the ability of serum antibody to induce activation of the complement cascade [42]. ADCD was induced by vaccination in both cohorts in response to only XBB.1.5. (**Figure 4A**). ADCD to ancestral and BA.5 S was high in pre-vaccination samples suggesting high preexisting immune responses through complement activation in both cohorts. Non-pregnant females had decreased post vaccination ADCD to JN.1, which was significantly lower than pregnant females which were unchanged by vaccination. These data highlight the differences between variants and how virus evolution may dictate the type and function of antibody generated in response to vaccination. XBB.1.5 and JN.1 are distinct lineages originating from BA.5 that harbor many mutations on the spike protein which can alter the ability of the virus to evade distinct immune functions [57]. While both variants have evolved antibody evasion strategies, these data suggest they may be leveraged through different mechanisms.

**Figure 4.**
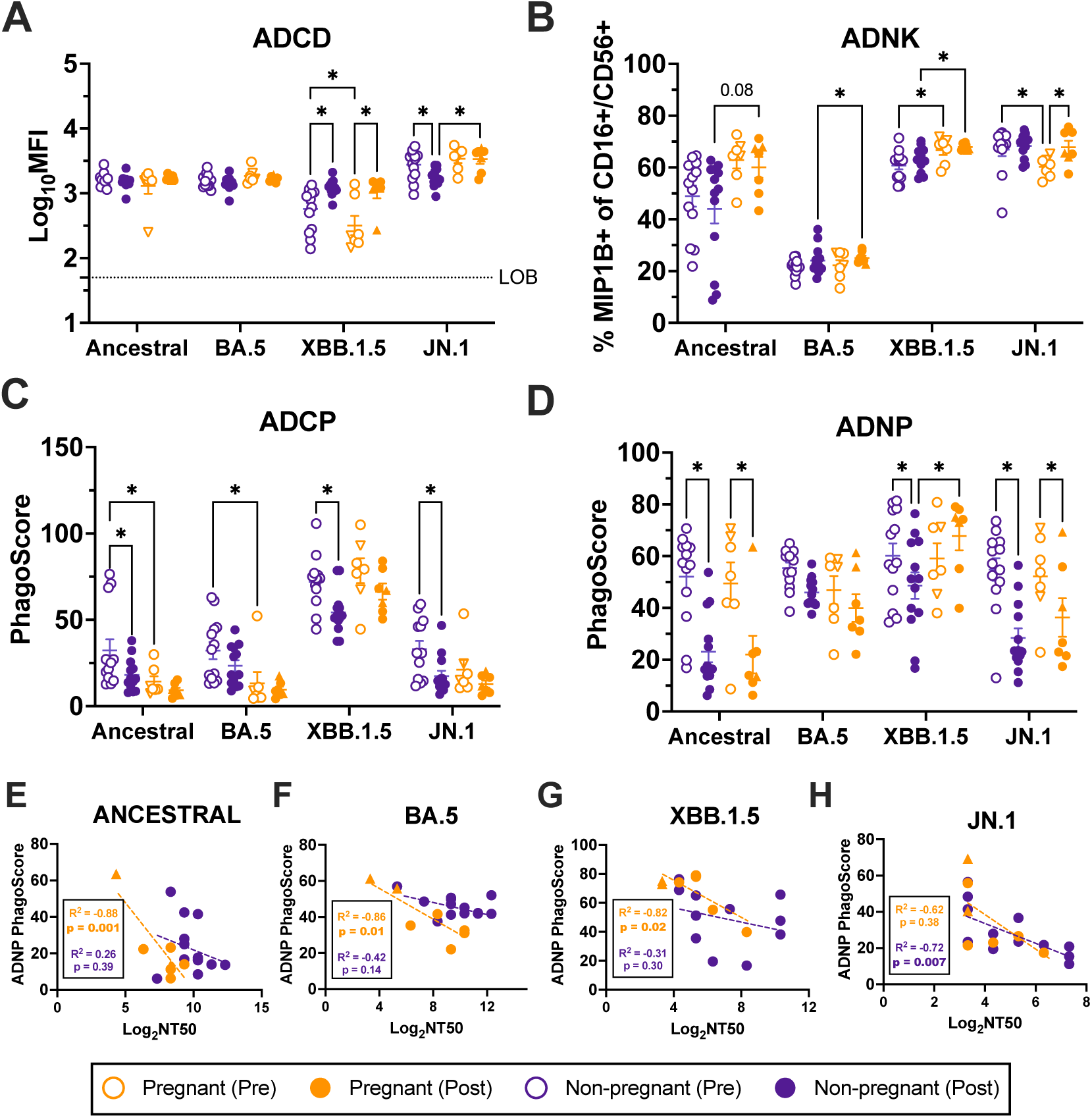
Non-neutralizing and neutralizing antibody functions are negatively correlated among pregnant women. Non-neutralizing antibody functions were measured pre and 3-5 weeks post-vaccination. Pregnant women (n=7) are shown in orange and non-pregnant females (n=13) in purple. Triangles highlight women who received monovalent boosters and circles represent bivalent booster recipients. Horizontal bars indicate mean MFI across samples with bars indicating standard error of the mean. **(A)** antibody dependent complement deposition (ADCD) was measured by assessing antibody binding to C3 protein and is reported as log_10_ mean fluorescence intensity. Stippled line indicates limit of background (LOB) defined as the log_10_MFI of a defined negative control. NK cells isolated from donor blood were utilized to assess **(B)** antibody dependent NK cell cytotoxicity (ADNK), or the ability of antigen specific serum antibody to include NK cell cytokine production or degranulation. Data are reported as % MIP1B+ cells of CD16+/CD56+ NK cells. **(C)** THP-1 monocyte cell line was used to measure antibody dependent cellular phagocytosis (ADCP), or the ability of antibody to induce phagocytosis of antigen. **(D)** Neutrophils isolated from donor blood were used to measure antibody dependent neutrophil phagocytosis (ADNP), or the ability of antibody to induce neutrophil phagocytosis of antigen. ADCP and ADNP are reported as a PhagoScore, calculated as (% bead positive monocytes/neutrophils) * (geometric MFI of each cell). **(E-H)** Pearson correlations between post-vaccination neutralizing antibody titers (Log_2_NT50) and ADNP PhagoScores for are shown for: **(E)** ancestral, **(F)** BA.5, **(G)** XBB.1.5, and **(H)** JN.1 viruses. (A-E) Data were analyzed by linear mixed effects regression analysis with Bonferroni post-hoc with asterisks indicating p < 0.05. (E-H) Separate Pearson correlation R^2^ and p-values are shown for each variant separated by pregnancy status, with p < 0.05 shown in bolded font. Regression lines for each group are shown in dashed lines colored according to cohort.

In pregnancy, macrophage inflammatory protein (MIP)-1β+ NK cells are found at the maternal-fetal interface and are important for successful implantation and placentation [58]. In NK cells, MIP-1β acts as a chemoattractant in response to activation [59]. Antibody-dependent NK-cell cytotoxicity (ADNK) can be measured through MIP-1β production by NK cells [60]. Although neither cohort induced additional MIP-1β secretion against ancestral, BA.5, or XBB.1.5 following vaccination, MIP-1β+ NK cells were uniquely elevated in pregnant women following vaccination when compared with non-pregnant females (**Figure 4B**). In NK cells, interferon gamma (IFNγ) acts as an antiviral cytokine to promote further accumulation, activation, and cytotoxicity of NK cells [61]. As an additional readout of ADNK, NK cell IFNγ production post-vaccination was elevated in pregnant women in response to BA.5 and in non-pregnant females in response to JN.1 **(Supplemental Figure 3A).** There were no changes post-vaccination in NK-cell degranulation, as marked by CD107a+ NK cells, among either cohort **(Supplemental Figure 3B)**. While vaccination did not increase ADNK capacity to vaccine antigens in either cohort, elevated ADNK in pregnant women in comparison to non-pregnant women may be important for understanding the antiviral and functional capacity of antibodies generated by vaccination.

Antibody-dependent cellular phagocytosis (ADCP) and antibody-dependent neutrophil phagocytosis (ADNP) assess the ability of antibody to induce macrophage and neutrophil phagocytosis of antigen, respectively, and are reported as a PhagoScore [43], representing both the function and number of beads phagocytosed. While inconsistent across variants and groups, ADCP tended to decrease among non-pregnant women while no changes in PhagoScores were observed in pregnant women. ADNP followed a similar trend among non-pregnant women. Among pregnant women, however, ADNP decreased against ancestral and JN.1 antigens while there was no change in response to either BA.5 or XBB.1.5 post vaccination (**Figure 4C**). This is likely indicative of shifting dynamics between neutralizing and non-neutralizing functions, a phenomenon that has been documented in the context of repeat vaccination and may be a result of immune imprinting mechanisms [62]. Correlation analyses revealed significant negative correlations between nAb titers and ADNP PhagoScores among pregnant women only, further suggesting a shift away from neutralization, and towards non-neutralizing neutrophil phagocytosis among pregnant women (**Figure 4E-H**). For JN.1, correlation analyses were non-significant likely because responses from many pregnant women were below the limit of detection for nAb titer (**Figure 4H**). In non-pregnant females, while negative correlations between nAb titers and ADNP were seen in response JN.1, ancestral, BA.5 and XBB.1.5, these associations were non-significant, suggesting small sample sizes may preclude strong conclusions.

### Altered correlations among IgG subclasses and antibody functional phenotypes in pregnant women

Correlation matrices can be used to examine complex relationships between antibody isotypes and effector functions assessed by systems serology [63]. These analyses can highlight isotype dependence and may reveal differences in subclass functionality [63]. Spearman correlation matrices were generated to assess correlations between IgG isotypes and FcR binding and antibody functional properties prior to and in response to vaccine and variant viruses (**Figure 5**). Prior to vaccination, there was no IgG subtype dominance, but there were stronger positive associations between IgG subclasses and antibody functional outcomes, including in response to variants (**Figure 5A)**. Matrices of post-vaccination samples were most different between pregnant and non-pregnant women for variants, with fewer strong correlations in pregnant than non-pregnant women. Among non-pregnant women, anti-S IgG1 was positively and anti-S IgG3 was negatively correlated with FcR binding as well as neutralizing and non-neutralizing antibody functions, particularly against variants. This trend was reversed in pregnant women, in which anti-S IgG3 was positively and anti-S IgG1tended to be negatively correlated with Fc receptor binding and functional outputs. These data further implicate the reduced IgG1:IgG3 ratio and isotype subclass switching as functionally altering antibody-mediated immunity in pregnant women.

**Figure 5.**
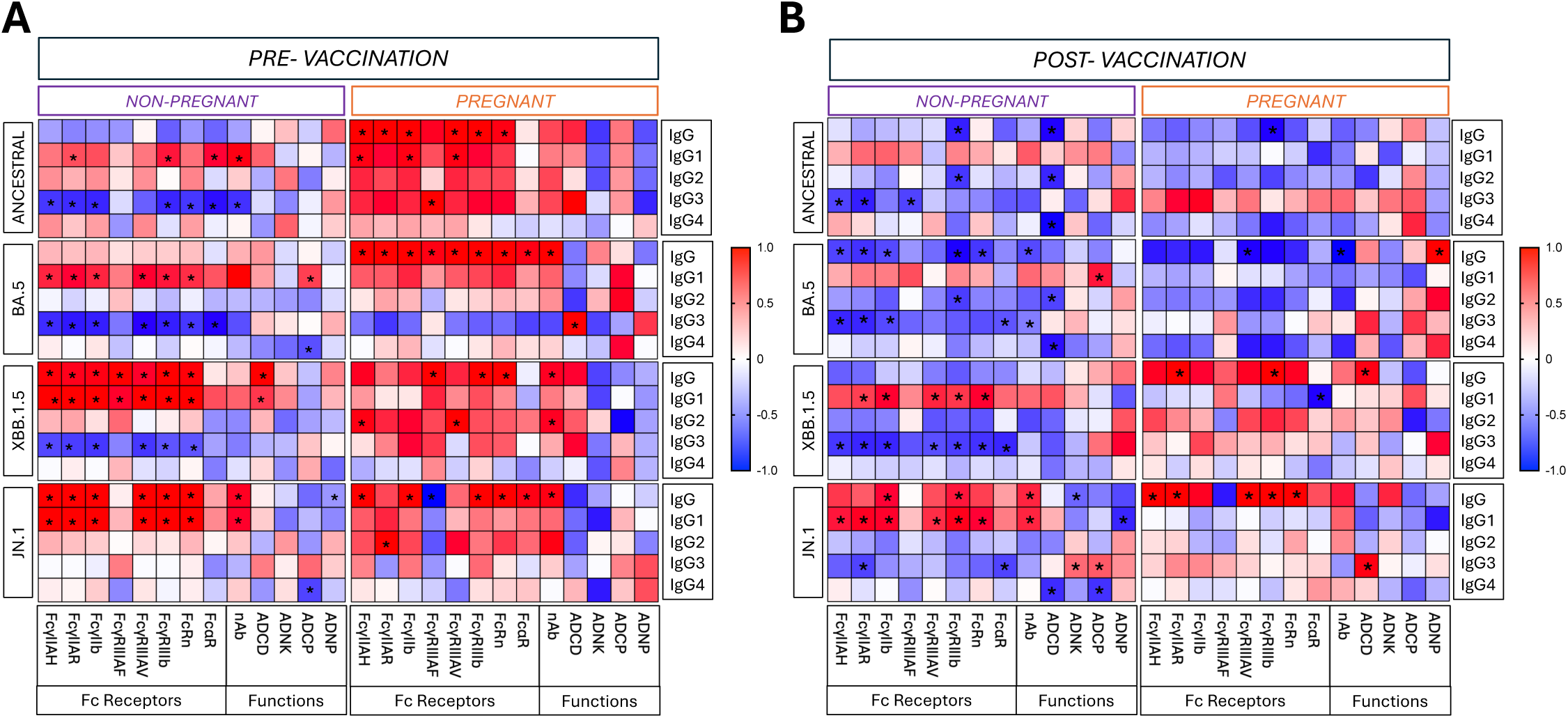
Correlations among IgG subclasses and antibody functional phenotypes differ between pregnant and nonpregnant women. **(A**) Pre-vaccination and **(B)** post-vaccination systems serology measures including isotype subclasses, FcR binding, non-neutralizing functions, and neutralizing antibody titers were log_10_ transformed and used for Spearman correlation analyses. Isotypes (y-axis) were correlated with functional measures (x-axis). Analyses were performed by pregnancy status. Asterisks indicate p< 0.05. Colors indicate Spearman correlation R value with a positive correlation of 1 indicated by deep red and a negative correlation of 1 indicated by deep blue. Abbreviations defined as followed: ADNP – antibody dependent neutrophil phagocytosis; ADCP – antibody dependent cellular phagocytosis; ADNK – antibody dependent NK cell cytotoxicity; ADCD – antibody dependent complement deposition; NT – neutralizing antibody titer.

### Decreased polyfunctional CD4^+^ T cells responses in pregnant women

Because immunoglobulin subtype switching depends on CD4^+^ T-cell help and cytokine production [64], we hypothesized that differences in CD4^+^ T-cell cytokine production between pregnant and nonpregnant females may contribute to differences in isotype switching and IgG subtypes. We assessed intracellular-cytokine production by memory CD4^+^ T cells **(Supplemental Figure 3A)** and CD8^+^ T cells **(Supplemental Figure 4A)** in pregnant and non-pregnant women in response to ancestral S-peptide pools [65, 66] prior to and after vaccination by measuring IFNγ, TNF, IL-2, and IL-21 secretion **(Supplemental Figures 3B, 4B)**. While individual cytokine frequencies provided initial insight into S-specific CD4^+^ memory T cell responses, comprehensive assessment of T cell functionality requires analysis of polyfunctional responses. Polyfunctionality is defined as the ability for individual cells to produce more than one cytokine simultaneously [67]. This approach is particularly important considering preliminary analysis of limited class-switching in pregnant participants, with polyfunctional CD4^+^ T-cell responses being known to enhance nAb titers and immunity following vaccination against multiple pathogens, including SARS-CoV-2 [68–70]. To characterize the functionality of CD4^+^ T cell responses, Boolean gating analysis identified all (15) possible combinations of the four measured cytokines represented as pie charts (**Figure 6A-B**) or histograms (**Supplemental Figure 5**). Intracellular cytokine staining of CD4^+^ T cells in response to ancestral S-peptide stimulation at post-vaccination timepoints suggested differences in cytokine profiles between groups **(Figure 6).** While both pregnant and non-pregnant women had IFNγ+ monofunctional CD4^+^ T cells, pregnant women tended to have proportionally higher frequencies of these cells. In CD4^+^ T cells, IFNγ production is known to modulate class switching in B cells, particularly favoring IgG3 over IgG1 [71–73]. Non-pregnant women showed a trend toward greater CD4^+^ T cell polyfunctionality, with more cells co-expressing multiple cytokines including IL-21, TNF and IL-2 (**Figure 6B**), cytokines that influence B cell maturation and antibody production. IL-2 is essential for T cell proliferation and survival, particularly supporting the development and maintenance of T follicular helper (Tfh) cells for germinal center reactions [74]. TNF contributes to germinal center formation and promotes B cell differentiation and isotype switching [75, 76]. Our analysis revealed that TNF production was enhanced in memory CD4^+^ T cells from pregnant but not non-pregnant females after vaccination. Alterations in polyfunctionality of T cells, and decreased production of these cytokines may contribute to the diminished cross-neutralizing antibody responses and altered IgG subclass distribution among pregnant women. In contrast, no effects of pregnancy or vaccination were observed in intracellular cytokine responses to S in memory CD8^+^ T cells (**Supplemental Figure 6**).

**Figure 6.**
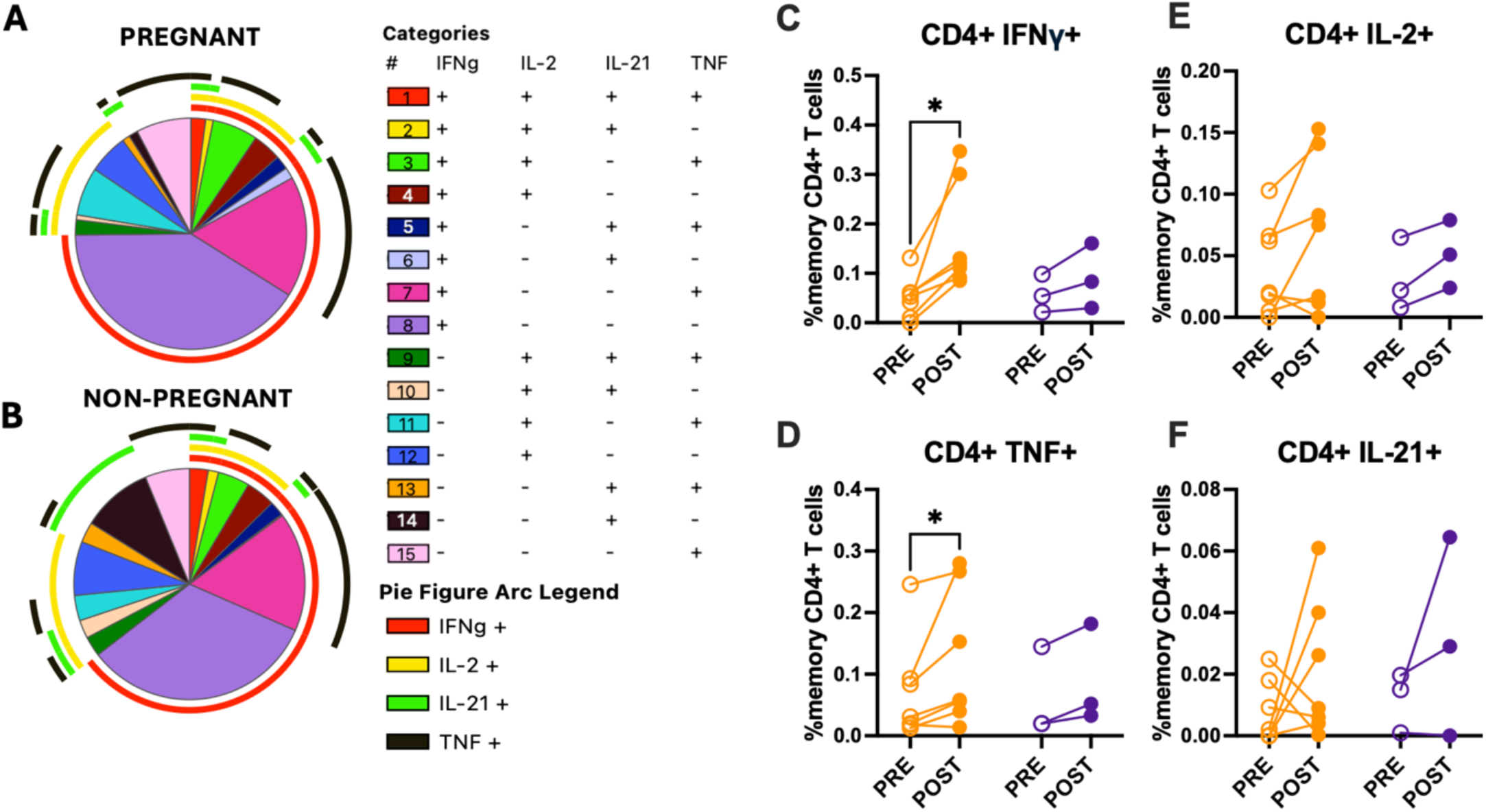
CD4+ T cells have reduced polyfunctionality among pregnant women. Peripheral blood mononuclear cells (PBMCs) were collected pre and 3-5 weeks post-vaccination for flow cytometry analyses. Cytokine-producing, antigen-specific CD4^+^ T cells for **(A)** pregnant (n=7) and **(B)** non-pregnant (n=3) women represented in pie charts broken down by the 15 cytokine combination categories. Arcs identify portions of the pie that express each specific cytokine. Paired dot plots demonstrate the percent of memory CD4^+^ T cell producing **(C)** IFNγ, **(D)** TNF, **(E)** IL-2, or **(F)** IL-21 following stimulation with ancestral SARS-CoV-2 spike peptide pools pre- and post-vaccination. Data were analyzed by linear mixed effects regression analysis with Bonferroni post-hoc. Asterisks indicate p< 0.05.

### Carnitine palmitoyltransferase 1A (CPT1a) expression is elevated *in* T cells of pregnant women

To further investigate phenotypic differences in T cells from pregnant and non-pregnant women, data generated by UMAP (Uniform Manifold Approximation and Projection) and clustering analysis were analyzed for shifts pre- and post-vaccination in both groups. There were no differences in any markers of activation, exhaustion, and metabolism (**Supplemental Table 1**) pre- or post-vaccination between pregnant and non-pregnant except for carnitine palmitoyl transferase 1 A (CPT1a), the rate limiting enzyme in long chain fatty acid oxidation (FAO) [77]. CPT1a was greater in both CD4^+^and CD8^+^ T cells prior to vaccination among pregnant compared with non-pregnant females (**Figure 7A-F**). Quantitative analysis of CPT1a mean fluorescence intensity [78] in memory CD4^+^ T cells revealed that pregnant females maintained approximately 1.5-fold higher expression (31,646 ±,1,372) compared to non-pregnant females (22,885 ± 1,587) prior to and after vaccination **(Figure 7A-B).** This was also seen in the pre-vaccination timepoint for CD8+ memory T cells **(Figure 7D).** This is consistent with prior literature showing that gestational changes increase fatty acid availability to T cells [79]. After vaccination CPT1a was reduced in total and memory CD4^+^ T cells (**Figure 7C-D**), but not in either total or memory CD8+ T cells (**Figure 7E-F**) among pregnant females. In contrast, CPT1a remained constant after vaccination among non-pregnant females.

**Figure 7.**
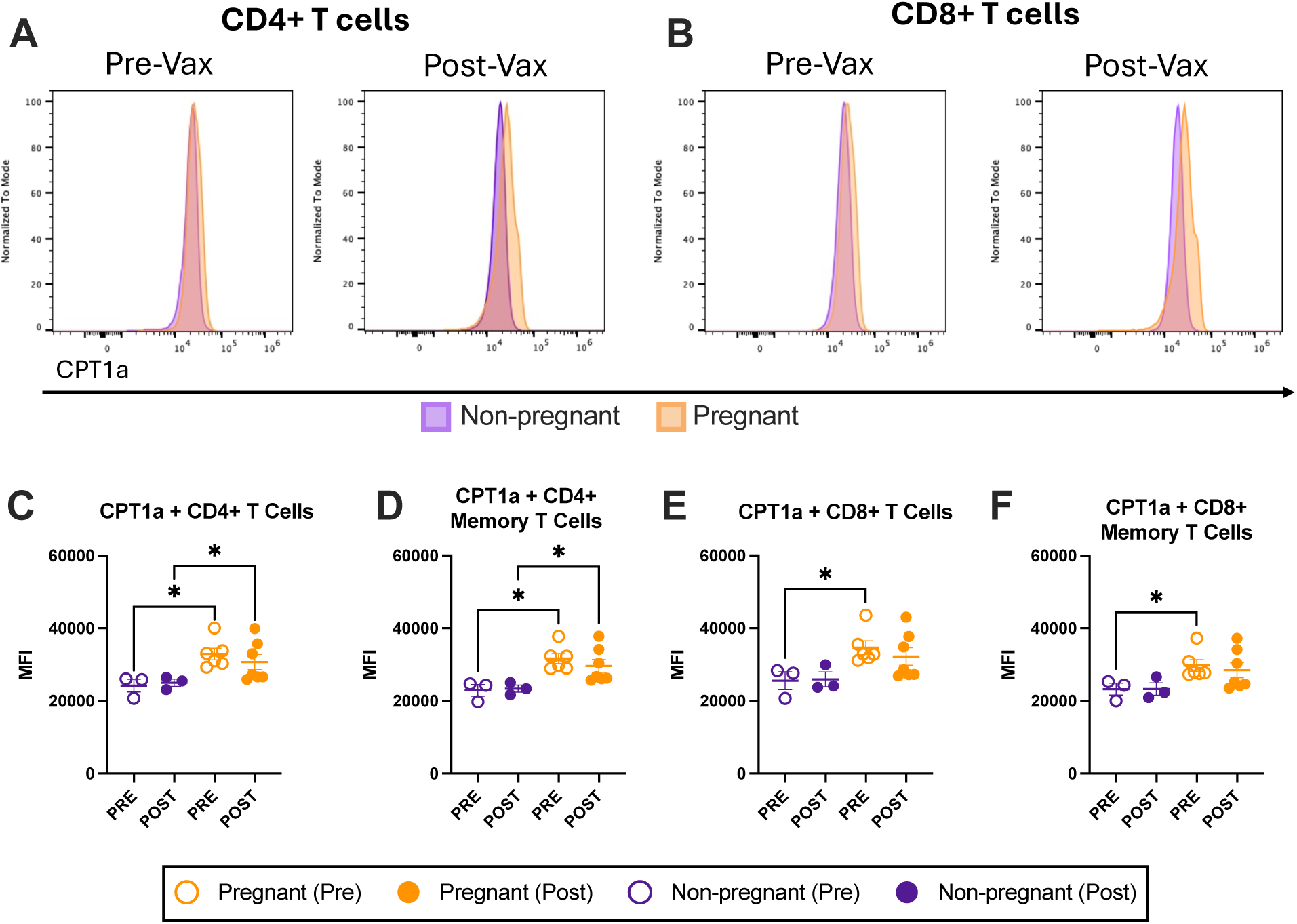
Carnitine palmitoyl transferase 1A (CPT1a) expression in CD4+ and CD8+ T cells pre- and post-vaccination is higher in pregnant women. Peripheral blood mononuclear cells (PBMCs) were collected pre and 3-5 weeks post-vaccination for flow cytometry analyses. Histograms showing CPT1a expression in total **(A)** CD4^+^ and **(C)** CD8^+^ T cells from peripheral blood mononuclear cells collected pre- and 3–5 weeks post-vaccination. Pregnant individuals (n=7) are shown in orange and non-pregnant individuals (n=3) in purple. Quantification of mean fluorescence intensity in memory CD4^+^ **(B)** and memory CD8+ **(D)** T cells are shown for each group pre- and post-vaccination. Horizontal bars indicate mean MFI across samples with bars indicating standard error of the mean. Data were analyzed by linear mixed effects regression analysis with Bonferroni post-hoc. Asterisks indicate p< 0.05.

## Discussion

Through broad characterization of the antibody landscape following COVID-19 vaccination, pregnancy was associated with reduced cross-reactive antibody responses, reflected in decreased IgG1:IgG3 ratios, nAb titers, and FcR-binding in pregnant compared with non-pregnant women. Previous studies have assessed the antibody profile generated by COVID-19 vaccination during pregnancy [31, 33, 80], with a focus on transplacental antibody transfer and protection of babies and infants from severe disease. One study showed limited cross-recognition in pregnant women to non-vaccine antigens after the first doses of ancestral vaccines, without comparison to non-pregnant controls nor assessment of neutralizing responses [31]. Our data illustrate that pregnancy is associated with reduced breadth and lower nAb responses to variants as compared with non-pregnant controls. Continuing efforts to assess antibody responses to updated vaccines remains critical to understanding cross-protection to variants in circulation.

SARS-CoV-2 has continued to rapidly evolve and mutate into new variants to escape adaptive-immune responses [81]. In 2022, Omicron variants became dominant in circulation, largely due to an ability to evade nAb generated by ancestral vaccines and natural infections [82]. New variants with the ability to evade nAb continue to emerge and dominate, with XBB.1.5. appearing in October 2022 and peaking in January of 2023, after the inclusion of BA.5 in bivalent booster vaccines was established [83]. By the time XBB.1.5 monovalent boosters were accessible, a new variant, JN.1 caused 60% of global cases [83, 84]. A potential limitation of annual vaccines is mismatches between dominant circulating strains and updated booster vaccines because variants continue to emerge between annual vaccines strain updates [85–87]. While mRNA technology has increased the speed in which companies can adapt updated vaccines to emerging variants, development of a new booster vaccine still takes several months [88]. Thus, government officials and researchers must work collaboratively to select strains for booster vaccines that will elicit the greatest breadth of immunity [88]. In our study, almost all women were vaccinated against ancestral and BA.5 (bivalent boosters), while XBB.1.5. was the dominant variant in circulation. Our data suggest that pregnancy may limit cross-protective antibodies.

Retrospective chart analysis shows that pregnant women are at an elevated risk for breakthrough infection [89]. Similar risk has been shown for other immunologically unique groups including solid-organ transplant recipients and those with autoimmune disease [89]. Thus, our findings of reduced cross-reactive and cross-neutralizing antibodies may provide insight into the mechanism driving elevated risk for breakthrough infection among vulnerable populations.

With repeated exposure to antigen through natural infection and booster vaccines, antibody responses to COVID-19 vaccines have evolved and correlates of protection have changed. Total IgG and nAb titers are considered adequate correlates of protection against COVID-19, similar to other acute respiratory infectious diseases [90]. Recent data have suggested that to mount sufficient protection, antibodies against SARS-CoV-2 must possess both neutralizing and non-neutralizing functions as correlates of protection [91]. As nAb titers wane and symptomatic cases rise, hospitalization rates among the general population remain low [92]. This provides evidence that other antibody characteristics may be important for protection from severe disease. Our data demonstrate that pregnant women have reduced FcR binding and nAb responses combined with elevated ADNK, especially to non-vaccine antigens.

Pregnancy is known to alter antibody glycosylation patterns which can dramatically affect their ability to efficiently and effectively interact with Fc receptors [93, 94]. Structural changes to the antibody-receptor interaction during pregnancy drives nuanced functional consequences that alter cross-reactivity and potentially cross-protection, generated in response to vaccination [93, 94]. In the context of SARS-CoV-2, non-neutralizing antibodies capable of inducing NK cell cytotoxicity were preferentially transferred across the placenta rather than nAb and was driven by differences in antibody glycosylation patterns [95].

Future work characterizing antibody glycosylation patterns in pregnant and non-pregnant women could shed light on the mechanisms by which pregnancy alters cross-reactive antibody responses.

Our data suggest that reduced class switching and T-cell polyfunctionality may contribute to limited cross-reactivity of antibodies during pregnancy. CD4^+^ T cells are critical for generation of high affinity, class switched antibodies and contribute to vaccine-induced protection against COVID-19 [96]. Expansion of IFNγ+ monofunctional CD4^+^ T cells may contribute to reduced IgG1:IgG3 ratios among pregnant women, as these cells may promote IgG3 bias in B cells [71–73]. The observation of a pregnancy-associated metabolic shift toward enhanced FAO compared to non-pregnant participants may reflect a complex dynamic of baseline metabolic reprogramming during pregnancy that may alter vaccination-induced immune responses. Maternal circulation provides a fatty acid-rich environment during the third trimester [79]. The increased expression of CPT1a supports the ability of T cells to use FAO as an energy source in pregnancy. FAO influences the differentiation of CD4^+^ T cells into different subsets, but the impact of increased FAO on polyfunctionality is not well understood [97, 98]. The small sample sizes in our T cell analyses (N=3 non-pregnant, N=7 pregnant) limit the generalizability of our findings and statistical power to detect meaningful differences. However, despite these small numbers, a statistically significant increase in T cell CPT1a expression provides additional insights into the impact of a fatty acid rich environment in pregnancy. Additional investigation into how this may impact CD4^+^ T cell polyfunctionality could provide mechanistic insights into pregnancy-associated immune adaptations and highlight the importance of larger studies to confirm these preliminary observations and establish clinical significance to better inform vaccine adherence in the context of pregnancy.

While not studied here, maternal antibody transfer is a critical aspect of vaccination during pregnancy, and our data may have implications in our understanding of the protection provided to neonates via cord blood. In fact, a study assessing newborn antibody found that the total number of maternal vaccines received in addition to the timing of the most recent vaccination was a strong predictor of nAb in neonates [99].

However, data on the cross-reactivity and cross-protection of these maternally transferred antibodies is lacking. Data from pregnant women and their neonates receiving the primary vaccination series suggest limited Omicron specific IgG and FcR binding [31]. This suggests our data may translate to reduced neonatal protection from maternal antibody transfer.

Prior immunization and infection history is an important contributor to immune responses following vaccination and was not analyzed here due to low sample size. In our cohort, there was large amounts of pre-existing immunity reflected by high pre-vaccination levels to a variety of measures including total IgG. Unfortunately, we were unable to directly assess the impact of prior exposures as vaccination and infection records were not available for either cohort of participants. As such it is important to consider the role of immune imprinting when considering cross-reactive and cross-protective antibodies generated through vaccination as immune history may direct the breadth and depth of antibody responses [62]. From data in non-pregnant individuals, immune imprinting is shown to significantly limit the ability of B cells to generate antibody responses to vaccine variants following primary vaccination series [100]. This is an important consideration, particularly as our data suggest pregnancy limits the cross-reactive antibody response and may be further influenced by immune imprinting mechanisms.

Although participants in the pregnant and non-pregnant cohorts were age-matched, small sample size limited our ability to consider other demographic variables and impacts the generalizability of our findings. For example, previous studies have shown the impact of trimester on immune responses generated by COVID-19 vaccination [16, 101]. In the present study, sample size limited our ability to assess trimester differences, with 6/7 pregnant females enrolled in our study being vaccinated in the second trimester of pregnancy. Additionally, while race is an important consideration to understand differences in immune responses to vaccination, small sample size has limited our ability to stratify data in this way. Consideration of these demographic factors, among others is important and should be a focus of future studies. The small sample size is a major limitation of this study and was associated with challenges recruiting pregnant women. While 232 pregnant women were approached during the study enrollment window, only 7 completed the study. This likely reflects the limited COVID-19 vaccine uptake among pregnant women, which we show is caused by numerous factors across the care continuum that contribute to COVID-19 vaccine hesitancy in this population. Identification of points of public-health intervention to effectively increase vaccine uptake among pregnant women is needed.

## Methods

### Study Design

Pregnant women were enrolled into mRNA vaccine studies at the Johns Hopkins Maternal & Fetal Medicine Clinic (IRB 00246472) (n=7 pregnant) during the 2022-2023 vaccination season. Pregnant women were contacted at clinical visits or via phone to determine interest and eligibility. Women were eligible if they planned to receive an mRNA COVID booster vaccine (Pfizer of Moderna) during weeks 32 through 36 of pregnancy, and consent to maternal blood collection before and after receipt of the booster vaccine. Patients who had already received an initial dose(s) of the vaccine were still eligible to enroll.

Participants were made aware of the study scope and the anonymity and confidentiality of their collected data. The voluntary nature of their participation, and the details of the study protocol were made evident. All participants gave informed consent to participate in the study. Throughout recruitment, 232 pregnant women were approached to participate in this study. Ninety-one patients were initially interested while only 14 enrolled in the study. Of these, 7 completed the study. Of these seven, two women (distinguished by triangles in all graphs) received monovalent ancestral boosters while the remaining five were recruited after the release of the bivalent ancestral BA.5 booster in August of 2023 and thus received this updated vaccine. For non-pregnant female comparators, samples were obtained from age-matched adult female healthcare workers who were recruited during the 2022-2023 influenza vaccine enrollment period by fliers, emails, and announcements about the annual vaccination program and were able to self-enroll upon receipt of the influenza vaccine at the Johns Hopkins East Baltimore hospital (IRB00288258) (n = 13 non-pregnant). All non-pregnant comparators selected as controls for this study received the bivalent ancestral BA.5 booster vaccine. While we have documentation on the vaccine type received at enrollment, prior vaccination and infection records were not available for pregnant nor non-pregnant participants. Demographic information is detailed in **Table 1**. Of the vaccinees enrolled, 13 age-matched non-pregnant females received the COVID-19 booster vaccine during the same vaccination season and had pre- and post-vaccination blood samples available. Plasma and peripheral blood mononuclear cells were obtained prior to and one month after receipt of mRNA booster vaccines from pregnant and non-pregnant females. Plasma was used to assess antigen-specific immunoglobulin isotype responses and neutralizing antibody responses, in addition to the crystallizable fragment (Fc) receptor binding and non-neutralizing antibody functions. Cryopreserved PBMCs were used for flow-cytometry analyses.

**Table 1.**

|  | Non-Pregnant<br>n = 10 | Pregnant<br>n = 7 | P-Value |
| --- | --- | --- | --- |
| **Demographics** | Mean ± SD | Mean ± SD | |
| **Age** | 31.8 ± 3.35 | 32.9 ± 3.18 | 0.5243734 |
| **Vaccine Type** | 0 0% | 2 28.5% |  |
| Monovalent Booster | 10 100% | 5 71.5% |  |
| Bivalent Booster |  |  |  |
| **Average Gestational Age (weeks)** | - | 22.4 ± 11.78 | |
|  | **N %** | **N %** |  |
| **Race** |  |  |  |
| White | 6 60.0% | 7 100% |  |
| Black | 2 20.0% | 0 0% |  |
| Asian | 2 20.0% | 0 0% |  |
| **Gestational Age (At Vaccination)** | - | 1<sup>st</sup> Trimester (n = 0) 0%<br>2<sup>nd</sup> Trimester (n = 6) 85.8%<br>3<sup>rd</sup> Trimester (n = 1) 14.2% |  |

### Cells and Viruses

VeroE6-TMPRSS2 cells were obtained from the cell repository of the National Institute of Infectious Diseases, Japan, and were grown in complete medium consisting of Dulbecco’s modified Eagle’s medium (DMEM) (Gibco, Thermo Fisher Scientific), containing 10% fetal bovine serum (FBS) (Gibco, Thermo Fisher Scientific), 1 mM glutamine (Invitrogen, Thermo Fisher Scientific), 1 mM sodium pyruvate (Invitrogen, Thermo Fisher Scientific), 100 U/mL penicillin (Invitrogen, Thermo Fisher Scientific), and 100 μg/mL streptomycin (Invitrogen, Thermo Fisher Scientific). Cells were incubated at 37°C in a humidified incubator with 5% CO_2_.

Ancestral (hCoV-19/USA/DC-HP00007/2020; B.1, GISAID EPI_ISL_434688), BA.5 (hCoV-19/USA/MD-HP32103-PIDCNSQVGY/2022, GISAID EPI_ISL_15013106), XBB.1.5 (hCoV-19/USA/MD-HP40900-PIDYSWHNUB/2022, GISAID EPI_ISL_16026423), and JN.1 (hCoV-19/USA/MD-HP49672-PIDDVSVURS/2023, GISAID EPI_ISL_18623285) variants of SARS-CoV-2 were isolated in VeroE6-TMPRSS2 cells plated in 24-well plates as previously described [35]. The consensus sequence of the virus isolate did not differ from the sequence derived from the clinical specimen. The infectious virus titer was determined on Vero-E6-TMPRSS2 cells using a 50% tissue culture infectious dose (TCID50) assay as described [36].

### Microneutralization assays

Plasma neutralizing antibodies [37] were determined as described for SARS-CoV and modified for SARS-CoV-2 as described [38]. Two-fold dilutions of plasma, starting at a 1:20 dilution, were made using incomplete medium (IM) - Dulbecco’s modified Eagle’s medium (DMEM) (Gibco, Thermo Fisher Scientific), containing 1 mM glutamine (Invitrogen, Thermo Fisher Scientific), 1 mM sodium pyruvate (Invitrogen, Thermo Fisher Scientific), 100 U/mL penicillin (Invitrogen, Thermo Fisher Scientific), and 100 μg/mL streptomycin (Invitrogen, Thermo Fisher Scientific). Infectious virus was added to the plasma dilutions at a final concentration of 1 × 10^3 TCID50/mL or 100 TCID50 per 100 μL. The plasma-virus solution was incubated at room temperature for 1 hour, and 100 μL of each dilution was added to 1 well of a 96-well plate of VeroE6-TMPRSS2 cells in hextuplicate. The cells were incubated for 6 hours at 37°C with 5% CO_2_. The inoculum was replaced with fresh IM, and the cells were incubated at 37°C with 5% CO_2_ for 3-4 days until cytopathic effect (CPE) is evident in the no serum controls. The cells were fixed by the addition of 100 μL of 4% formaldehyde for at least 4 hours at room temperature and then stained with naphthol blue-black overnight. The nAb titer was calculated as the highest serum dilution that eliminated the CPE in 50% of the wells (NT50), and the AUC was calculated [39, 40].

### Antigen-bead coupling

Antigen-specific isotyping and Fc-receptor binding were multiplexed using a Luminex Intelliflex in a 384-well format. Antigens used in this panel are as follows: ancestral spike (S) and nucleocapsid (N) protein, BA.5, XBB.1.5, and JN.1 spike proteins. These were obtained through the National Cancer Institute Serological Sciences Networks (SeroNet) for COVID-19 [41]. Each antigen was coupled to unique MagPlex bead regions (Luminex). Briefly, beads were activated by combining activation buffer (0.1M NaH2PO4; pH 6.2), 50mg/mL 1-ethyl-3-[3-dimethylaminopropyl] carbodiimide (EDC), and 50mg/mL Sulfo-NHS (N-hydroxysulfosuccinimide). Activated beads were coupled to 25µg of antigen in coupling buffer (0.05M MES; pH 5.0). Activated coupled beads were blocked with blocking buffer (PBS 1X, 0.1% BSA, 0.02% Tween-20, 0.05% sodium azide; pH 7.4), washed with washing buffer (PBS 1X, 0.05% Tween-20) and stored at 4°C in PBS.

### Luminex serological assays

Serum was diluted 1:25 and was incubated with bead solution (20 µL of each bead region in 20 mL of Luminex Assay Buffer: PBS 1X, 0.1% BSA, 0.05% Tween-20). This protocol was followed for isotyping (IgM, IgG [IgG1-4], and IgA [IgA1-2]) and binding to the following Fc receptors (FcRs): FcγRIIA-R, FcγRIIA-H, FcγRIIb, FcγRIIIA-V, FcγRIIIA-F, FcγRIIIB, FcRn, and FcαR (Duke Vaccine Institute – Protein Production facility). Following incubation, plates were placed on a magnet and washed with Procartaplex Wash Buffer (ThermoFisher). Each assay plate was then incubated with 45 µL of secondary antibody diluted in Luminex Assay Buffer. Isotypes were identified by PE-conjugated antibodies (Southern Biotech) and FcR proteins were biotinylated using BirA biotin-ligation kit (Avidity) at least one day prior and coupled to PE-Streptavidin (BioLegend) for 15 min on a rotor before use. Following incubation with secondary antibody, plates were placed on a magnet and washed. Beads were resuspended in Procartaplex Wash Buffer before running on the Luminex Intelliflex. Samples were run on FlexMap 3D Low with a DD range of 7000-1700.

For each assay, a minimum of 100 events was recorded per antigen and the MFI of secondary antibody-conjugated to PE, or FcR protein-conjugated to PE for each bead region was measured to quantify antigen-specific immunoglobulin isotypes and FcR responses in each sample at 0- and 28-days post vaccination [42]. LOB (limit of background) for these assays was set by PBS and IgG depleted sera as a negative control. Additional positive controls were included to ensure assay integrity. Optimal serum dilution was determined by a dilution curve to ensure all values fell within the limit of detection.

### Antibody-dependent complement deposition (ADCD)

Serum (1:15 dilution) was added to MagPlex antigen coated beads. Plates were incubated, placed on a magnet, and washed 3 times with 1% BSA. Lyophilized Low-Tox Guinea Pig Complement (Cedarlane) was reconstituted in ice-cold dH2O and diluted 1:25 in Gelatin Veronal buffer with Mg^2+^ & Ca^2+^ (GVB++) (Boston Bio). Diluted complement protein was added to each well followed by incubation at 37C. IgG goat anti-guinea pig C3 (MP Biomedicals) was diluted 1:500 in 0.1% BSA and added to each well. After incubation, Donkey-anti goat IgG secondary antibody-PE (ThermoFisher) was diluted 1:200 in 0.1% BSA and added to each well. Beads were resuspended in 0.1% BSA and run on the Luminex Intelliflex and data was collected as described above.

### Antibody-dependent NK-cell cytotoxicity (ADNK)

Natural killer (NK) cells were isolated from donor blood. Whole blood was centrifuged through a Ficoll gradient to harvest PBMCs and NK cells were isolated from PBMCs using the MagniSort™ Human NK-cell Enrichment Kit (ThermoFisher). Cell concentration was normalized to 1.5 x 10^6 cells/mL in R10 medium, and IL-15 (1 ng/mL, Abcam) was added and incubated overnight at 37°C. Plates were coated with 3 mg/mL antigen (ancestral, BA.5, XBB.1.5, and JN.1 spike proteins – SeroNet), blocked with 5% BSA, washed, and dried before adding serum (1:100 dilution). Inhibitor cocktail (2.5 µL CD107a-PeCy5 (BD Bioscience - BDB555802) + 0.5 mL BFA (Millipore Sigma) + 1 µL Golgi Stop (BD Bioscience) + 9 mL R10 medium) was added to each well, incubated, and surface staining cocktail (1 µL CD3-Pacific Blue (BD Bioscience - BDB558117) + 1mL CD56-PE-Cy7 (BD Bioscience - BDB557747) + 1 mL CD16-APC-Cy7 (BD Bioscience - BDB557758) + 7 mL PBS) is added to a V-bottom 96-well plate. NK cells were transferred to the V-bottom 96 well plate with pre-added surface stain cocktail and incubated. Cells were pelleted, supernatant removed, and Perm A (ThermoFisher) was added to each well. Cells were washed and Intracellular staining cocktail (1 mL IFNγ-APC (BD Bioscience -) + 1 mL MIP1-B-PE (BD Bioscience -BDB550078) + 48 mL Perm B (ThermoFisher) was added to each well. Plates were washed, cells were resuspended in PBS and run on a Symphony A3 through the Bloomberg Flow Cytometry and Immunology Core. Gating strategy as follows: NK cells-CD3-, CD16+, CD56+ with reported cell populations including IFNγ+, CD107a+ and MIP-1β + NK cells.

### Antibody-dependent neutrophil phagocytosis (ADNP)

Antigens (ancestral, BA.5, XBB.1.5, and JN.1 spike proteins – SeroNet) were biotinylated with the BirA biotin-ligation kit (Avidity) and coupled to FluoSpheres™ NeutrAvidin™-Labeled Microspheres, 1.0 μm, yellow-green fluorescent (505/515) (ThermoFisher). Antigen was combined with NeutrAvidin beads and incubated. Serum was diluted 1:100 in PBS and incubated with antigen coated beads. During incubation, cells were isolated from donor blood. Whole blood was collected into CPT tubes and centrifuged at 1000 rpm for 30 min. Granulocytes were collected from below the density gradient. Cells were suspended 1:10 in ACK Lysis buffer and incubated at RT for 5 min to lyse red blood cells. Cells were washed and resuspended in 10 mL of PBS. Plates were washed with PBS, 200µL of neutrophil solution (5 x 10^6 cells/plate) was added to each well, incubated, spun at 500g for 5 min, and cells were resuspended in extracellular staining solution (0.5µL APC CD66b (BioLegend - 305117) + 19.5mL PBS / well) and incubated for 15min covered with foil at RT. After staining, plates were washed and resuspended in 4% PFA to fix cells, washed and resuspended in PBS. Plates were run on the Attune NxT flow cytometer. Gating on CD66b+ cells (neutrophils) followed by bead+ neutrophils, stained with APC. Data are reported as a PhagoScore, calculated as (% bead-positive neutrophils) * (geometric MFI of each cell) [43].

### Antibody-dependent cellular phagocytosis (ADCP)

THP-1 cells, a human monocytic cell line, were grown in suspension and diluted with warm R10 medium. Following immune complex formation, assay plates are washed with PBS and centrifuged at 2000g for 10 min. The THP-1 cells were plated 2.5 x 10^4^ per well, and incubated O/N at 37°C. THP-1 cells were fixed, washed, and resuspended in PBS. Plates are run on the Attune NxT flow cytometer. Gating included bead+ single-cells stained with APC, representing THP-1 cells that have phagocytosed bead. Data are reported as a PhagoScore, calculated as (% bead-positive THP-1) * (geometric MFI of each cell) [44].

### Ex vivo flow cytometry staining

Intracellular cytokine-flow cytometry staining was used to assess antigen-specific cytokine production and T-cell phenotypic and functional profiles. Cryopreserved PBMCs were thawed, rested, and cultured overnight with 1 mg/mL peptide pools consisting of 315 peptides (15-mers with 11 amino acid overlap) spanning the ancestral SARS-CoV-2 spike protein (JPT Peptide Technologies, PM-WCPV-S-1) or a DMSO (unstimulated) control. After overnight incubation, cells were washed and stained for viability with Invitrogen Live/Dead Fixable Blue Dead Cell Stain (L34962) and Fc Block (564219). Cell-surface staining was performed with our T-cell panel surface-antibody cocktail mix (**Supplementary Methods Table 1A).** Cells were fixed and permeabilized with a 1X Fixation/Permeabilization buffer from eBioscience (FoxP3/Transcription Factor Staining Buffer Set). Intracellular staining was performed with intracellular cytokine-antibody cocktail mix (**Supplementary Methods Table 1B**). Cells were resuspended in 1% paraformaldehyde and run on the 4-laser (16UV-16V-15B-8R) Cytek Biosciences Aurora spectral flow cytometer. For ICS, data were processed using biexponential transformation and exported for further analysis in FlowJo to assess cytokine-producing, antigen-specific T cells (**Supplemental Figure 3-4**).

Polyfunctionality of these T cells was analyzed with Pestle v2.0 and SPICE 6 v6.1. Metabolic, activation, and exhaustions markers were analyzed via Uniform Manifold Approximation and Projection (UMAP) (v4.1.1), Xshift (v1.4.1), and ClusterExplorer (v1.7.6).

### Statistical analyses

GraphPad Prism v10.1 was used to perform univariate analyses with details about statistical tests used for each dataset described in further detail in figure legends.

Spearman correlations of systems serology measures were visualized in heatmaps generated using R v4.2.3 and version 1.0.12 of the package “pheatmap” (https://cran.r-project.org/web/packages/pheatmap [45]. For ADNP and ADNK assays, data were averaged across two healthy donors. Flow FCS files were analyzed using FlowJo v10 (10.8.1) software. Gating strategy for ICS can be found in **Supplementary Figures 3** and 4. ICS data were exported from Pestle v2.0 and SPICE 6 v6.1 for further statistical analyses detailed in figure legends.

## Data Availability

All anonymized data that support the findings of this study can be made available upon request from the corresponding author.

## Data Availability

All data produced in the present work are contained in the manuscript.

## Acknowledgements

We thank the clinical coordinators and vaccinees for making this study possible. We are especially grateful to Kimberly Jones-Beatty and Michelle Ufua for their assistance with recruitment. Thank you to Galit Alter, Ryan McNamara, and Taras Chicz for graciously training lab members and sharing protocols to develop systems serology methods. We thank Prakash Srinivasan for the use of the Attune Nxt flow cytometer and the Bloomberg Flow Cytometry and Immunology Core. This study was supported by the Food and Drug Administration (FDA) of the U.S. Department of Health and Human Services (HHS) as part of a financial assistance award [Center of Excellence in Regulatory Science and Innovation grant to Johns Hopkins University, U01FD005946] totaling $415,256 with 75% percentage funded by FDA/ OWH. The contents are those of the author(s) and do not necessarily represent the official views of, nor an endorsement, by FDA/OWH, or the U.S. Government. FDA CERSI U01FD005942 (SLK), NIH NIAID 75N93021C00045 (AP, RR), NIH NCI U54 CA260492 (SLK, ALC).

## Author Contributions

SLK, IB, AP, and ALC conceptualized the experimental design and group comparisons for this study. MAP, H-SP, JSL, LStC, PS, IB, JS, KF, RR, and ALC identified, acquired, stored, and distributed patient samples and clinical and demographic data for analyses. MAP, H-SP, PC, AW and JSL developed and validated serological assays. MAP, JS, LG and H-SP processed samples and completed technical assays. MAP, KR, and AY conducted statistical analyses. MAP, LG, KR, ALC, and SLK developed all tables and figures. MAP, ALC, SLK wrote the initial draft of the manuscript. MAP, JS, LG, HSP, AY, KR, PC, JSL, LAS, AW, CPG, LB, HG, PS, KF, RR, IB, JS, ALC, AP, and SLC provided substantive edits and approved the final draft of the manuscript.

