## Supplemental Figures for "Pregnancy Reduces COVID-19 Vaccine Immunity Against Novel Variants"

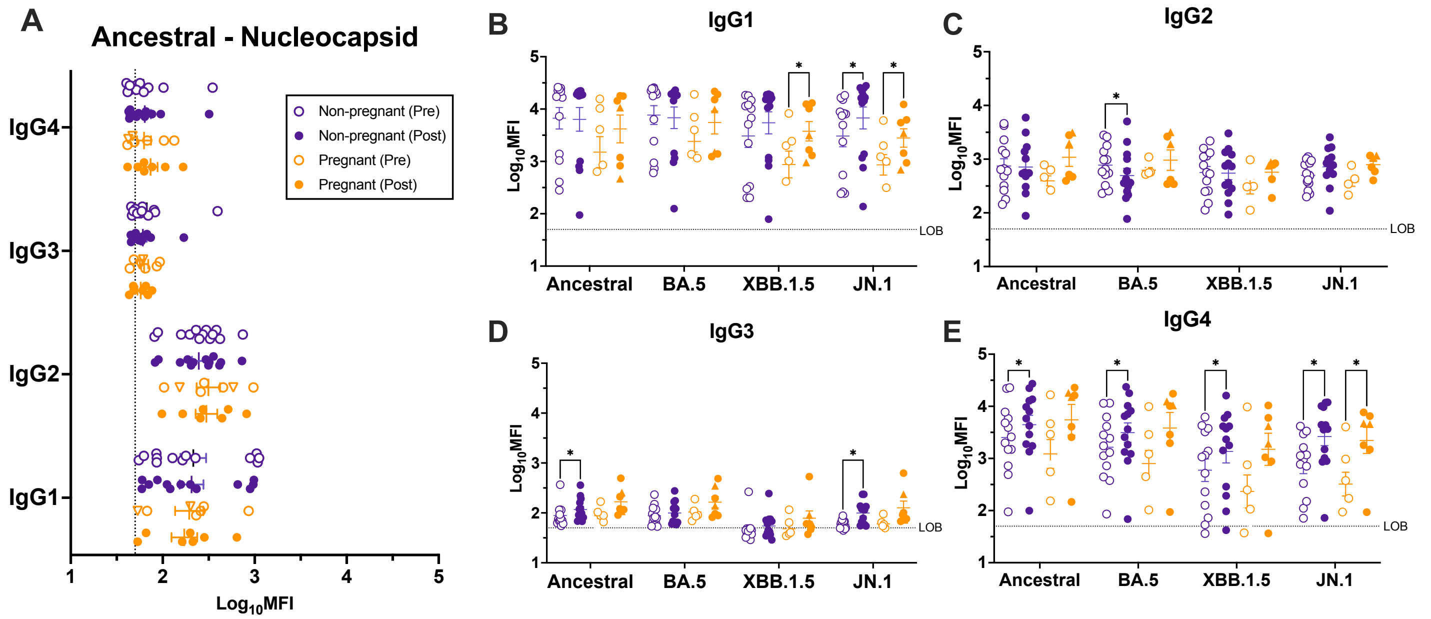


**Supplemental Figure 1. Vaccination differently induces IgG subtype responses during pregnancy.** IgG antibody responses in serum pre and 3-5 weeks post-vaccination were measured by systems serology and reported as $\log_{10}$ mean fluorescence intensity (MFI). Pregnant women (n=7) are shown in orange and non-pregnant females (n=13) in purple. Triangles highlight women who received monovalent boosters and circles represent bivalent booster recipients. Horizontal bars indicate mean MFI across samples with bars indicating standard error of the mean (SEM). **(A)** IgG subtype abundance to ancestral nucleocapsid (N) protein**.** IgG subtypes, including **(B)** IgG1 **(C)** IgG2, **(D)** IgG3, and **(E)** IgG4, in response to ancestral, BA.5, XBB.1.5, and JN.1 spike protein. Data were analyzed by linear mixed effects regression analysis with Bonferroni post-hoc. Asterisks indicate $p < 0.05$.

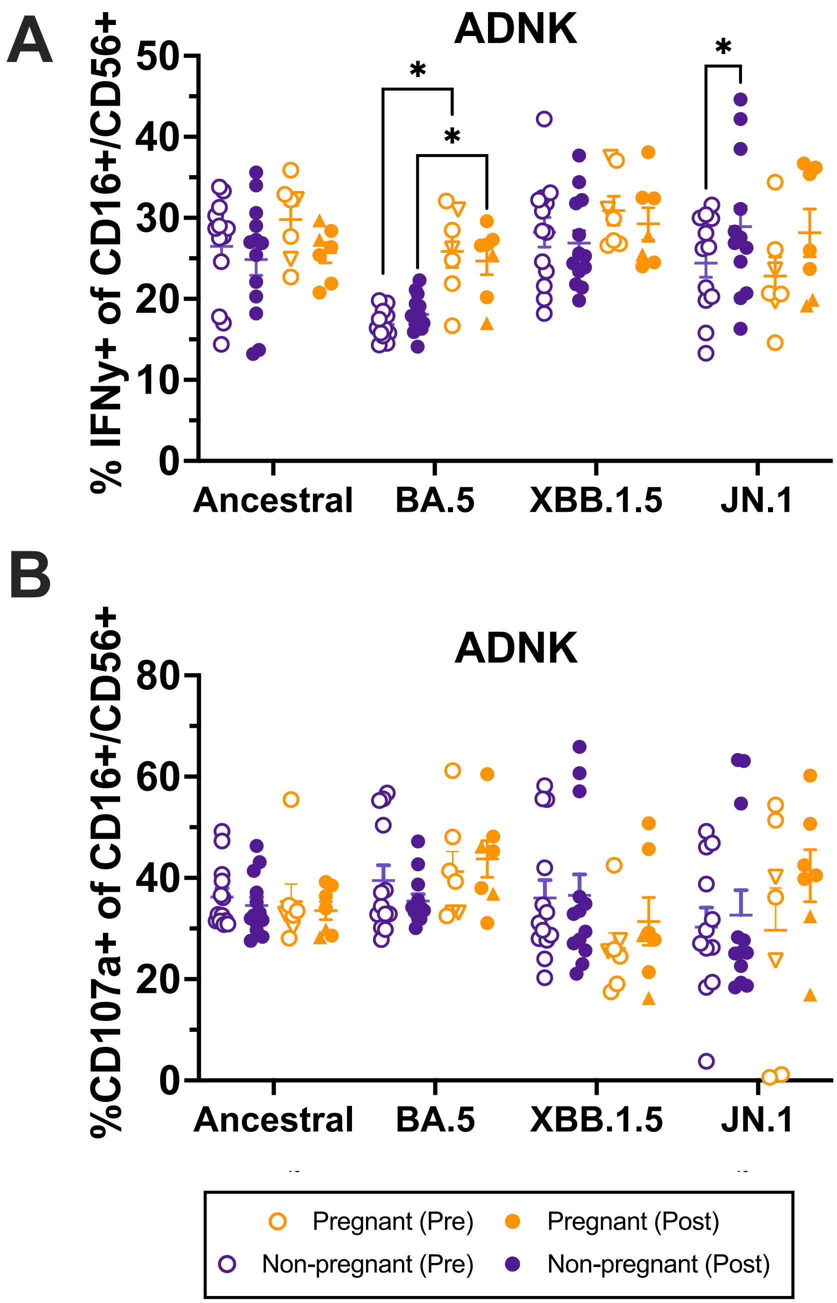


**Supplemental Figure 2. Additional antibody-dependent NK cell responses by pregnancy status.** NK cells isolated from donor blood were utilized to assess antibody dependent NK cell cytotoxicity (ADNK), or the ability of antigen specific serum antibody to include NK cell cytokine production or degranulation in serum pre and 3-5 weeks post-vaccination. Pregnant women (n=7) are shown in orange and non-pregnant females (n=13) in purple. Triangles highlight women who received monovalent boosters and circles represent bivalent booster recipients. Horizontal bars indicate mean MFI across samples with bars indicating standard error of the mean (SEM). ADNK was measured as ability to induce **(A)** IFNy production or **(B)** NK cell degranulation, as measured by CD107a positivity. Data were analyzed by linear mixed effects regression analysis with Bonferroni post-hoc. Asterisks indicate $p < 0.05$.

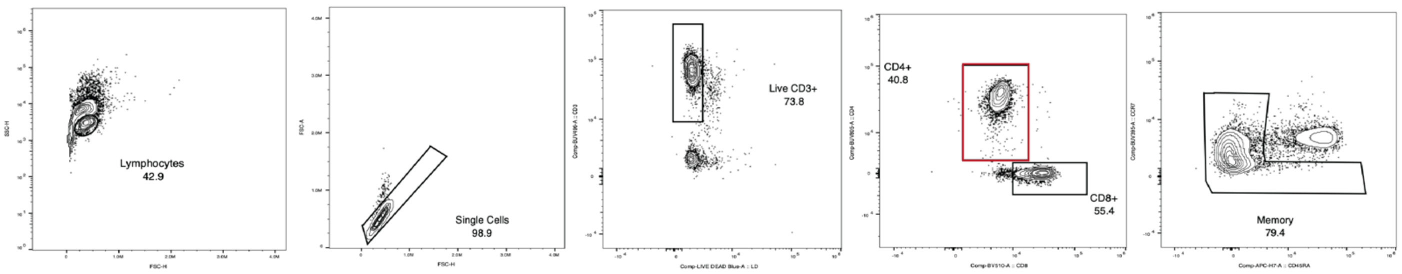


**Supplemental Figure 3A. Gating strategy for CD4⁺ memory T cells.** Representative flow cytometry gating of spike-specific memory CD4+ T cells

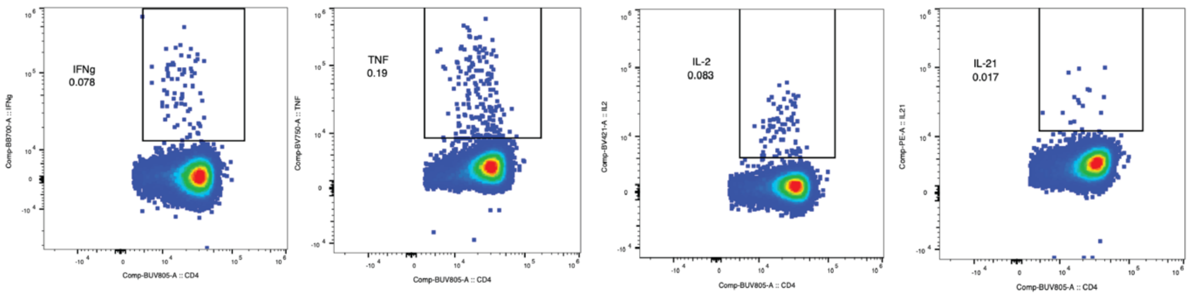


**Supplemental Figure 3B. Cytokine positive CD4⁺ T cells.** Representative flow cytometry gating of cytokine (IFNg, TNF, IL-2, and IL-21) producing spike-specific memory CD4+ T cells.

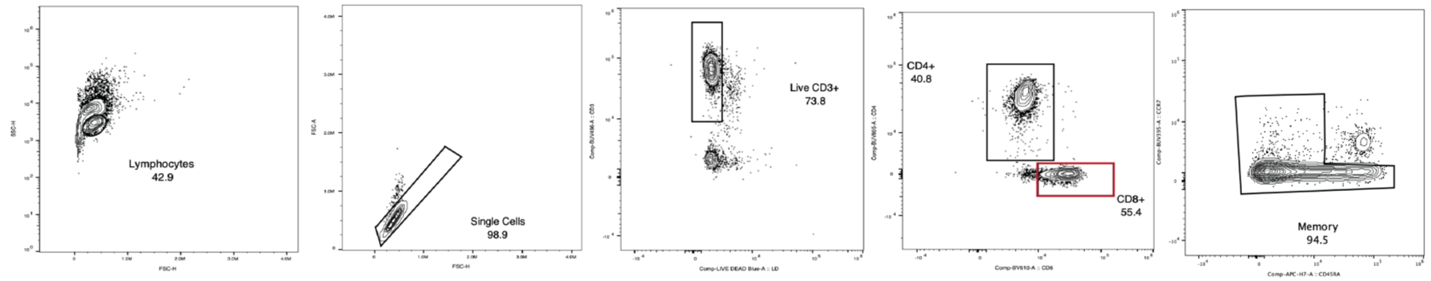


**Supplemental Figure 4A. Gating strategy for CD8⁺ memory T cells.** Representative flow cytometry gating of spike-specific memory CD8+ T cells

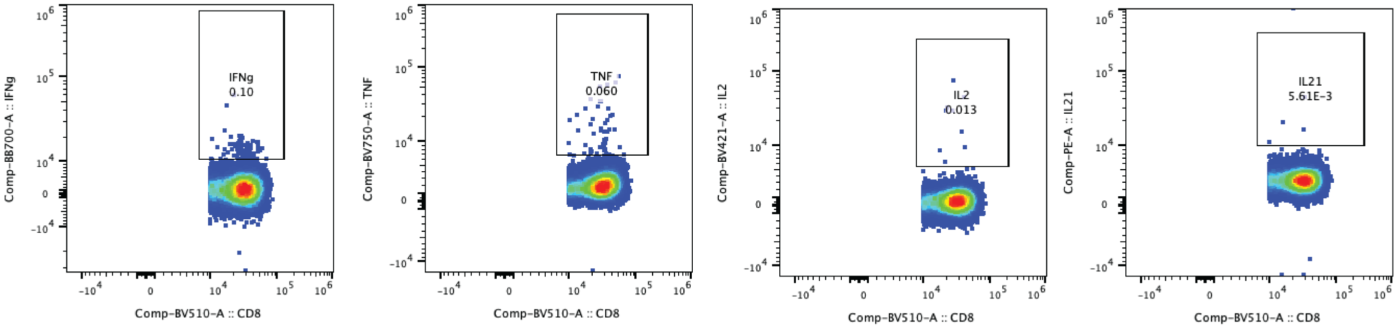


**Supplemental Figure 4B. Cytokine positive CD8⁺ T cells.** Representative flow cytometry gating of cytokine (IFNg, TNFa, IL-2, and IL-21) producing spike-specific memory CD8+ T cells.

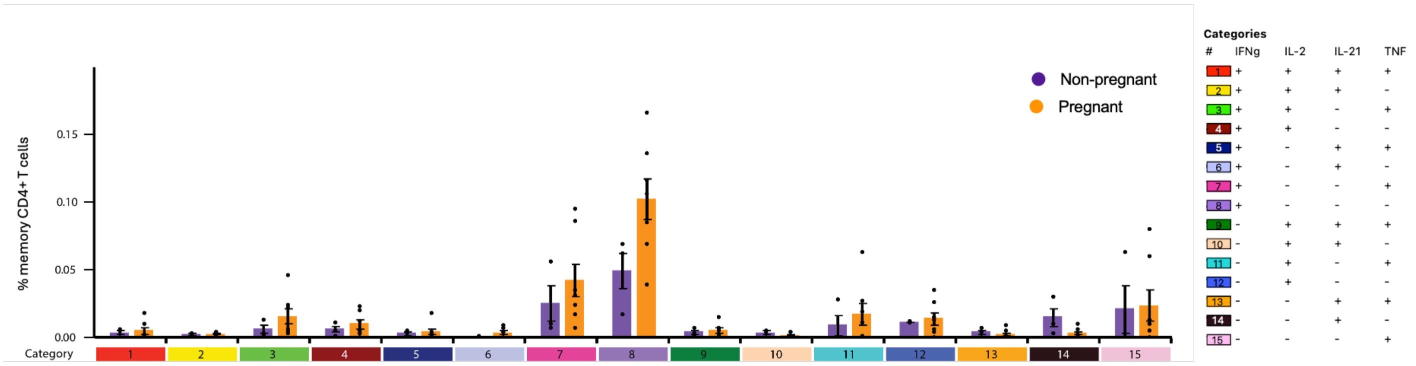


**Supplemental Figure 5. Pre- vs post-vaccination CD8+ T cells in pregnant and non-pregnant women.** Peripheral blood mononuclear cells (PBMCs) were collected pre and 3-5 weeks post-vaccination for flow cytometry analyses. Cytokine-producing, antigen-specific CD4+T cells for **(A)** pregnant (n=7) and **(B)** non-pregnant (n=3) women by the 15 cytokine combination categories. Histograms show the percent of memory CD4+ T cell producing cytokines following stimulation with ancestral SARS-CoV-2 spike peptide pools post-vaccination. Bars reflect mean values with standard error of mean.

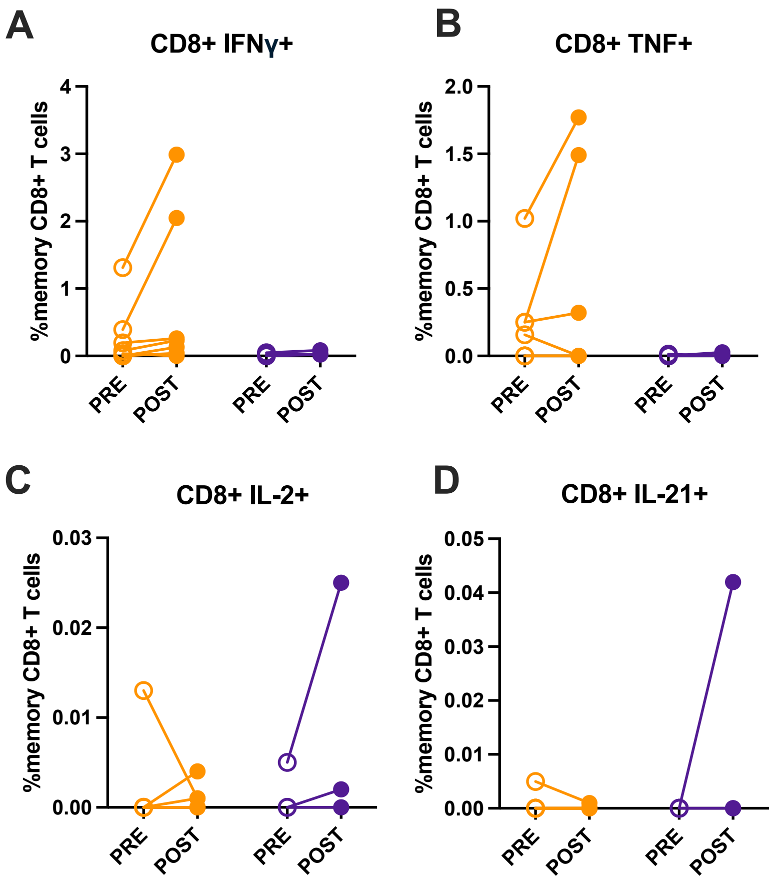


**Supplemental Figure 6. Pre- vs post-vaccination CD8+ T cells in pregnant and non-pregnant women.** Peripheral blood mononuclear cells (PBMCs) were collected pre and 3-5 weeks post-vaccination for flow cytometry analyses. Paired dot plots demonstrate the percent of memory CD8+ T cell producing (A) IFNγ, (B) TNF, (C) IL-2, or (D) IL-21 following stimulation with ancestral SARS-CoV-2 spike peptide pools pre- and post-vaccination.

**Supplemental Table 1. Intracellular cytokine staining (ICS) flow cytometry panel. (A)** surface staining markers and **(B)** intracellular markers.

**A. Surface**

| Fluorophore | Marker | Clone | Catalog | Vendor |
|---|---|---|---|---|
| BUV395 | CCR7 | 2-L1-A | 749655 | BD Biosciences |
| BV650 | CXCR3 | 1C6 | 740603 | BD Biosciences |
| PE CF594 | KLRG1 | 2F1 | 565393 | BD Biosciences |
| BV605 | CD69 | FN50 | 562989 | BD Biosciences |
| BUV563 | CD25 | 2A3 | 612918 | BD Biosciences |
| BUV661 | PD-1 | EH12.1 | 750260 | BD Biosciences |
| BUV737 | TIM-3 | 7D3 | 748820 | BD Biosciences |
| BV570 | CD28 | CD28.2 | 302926 | Biolegend |
| PE Cy5 | CD127 | HIL-7R-M21 | BD Custom | BD Biosciences |
| BUV805 | CD4 | SK3 | 612887 | BD Biosciences |
| APC H7 | CD45RA | HI100 | 560674 | BD Biosciences |
| BB660 | Tigit | 741182 | BD Custom | BD Biosciences |
| BV711 | OX40 | ACT35 | 563664 | BD Biosciences |
| BV786 | CD27 | L128 | 563327 | BD Biosciences |
| PE Cy7 | CTLA-4 | BNI3 | BD Custom | BD Biosciences |
| BV510 | CD8a | HIT8a | 300934 | Biolegend |
| BB790 | CXCR5 | RF8B2 | BD Custom | BD Biosciences |

**B. Intracellular**

| Fluorophore | Marker | Clone | Catalog | Vendor |
|---|---|---|---|---|
| PE | IL-21 | 3A3-N2 | 513004 | BioLegend |
| AF680** | HK2 | EPR20839 | ab228819 | Abcam |
| AF488 | TCF1 | 812145 | IC8224G | FisherScientific |
| BV421 | IL-2 | MQ1-17H12 | 564164 | BD Biosciences |
| AF532*** | VDAC1 | 20B12AF2 | ab14734 | Abcam |
| BV750 | TNF | MAb11 | 566359 | BD Biosciences |
| PECy5.5* | CPT1a | 8F6AE9 | ab128568 | Abcam |
| AF405 | Tomm20 | EPR15581-54 | ab210047 | Abcam |
| Alexa647 | GLUT1 | EPR3915 | ab195020 | Abcam |
| BUV496 | CD3 | UCHT1 | 612940 | BD Biosciences |
| BB700 | IFNy | B27 | 566394 | BD Biosciences |

Notes: * conjugated using PE/Cy5.5 Conjugation Kit - Lightning-Link (ab102899)

** conjugated using DyLight 680 Conjugation Kit (Fast) - Lightning-Link (ab201804)

*** conjugated using Alexa Fluor 532 Antibody Labeling Kit (Thermo Fisher, A20182)
